# Assessing the lack of diversity in genetics research across neurodegenerative diseases: a systematic review of the GWAS Catalog and literature

**DOI:** 10.1101/2024.01.08.24301007

**Authors:** Caroline Jonson, Kristin S. Levine, Julie Lake, Linnea Hertslet, Lietsel Jones, Dhairya Patel, Jeff Kim, Sara Bandres-Ciga, Nancy Terry, Ignacio F. Mata, Cornelis Blauwendraat, Andrew B. Singleton, Mike A. Nalls, Jennifer S. Yokoyama, Hampton L. Leonard

## Abstract

**Importance:** The under-representation of participants with non-European ancestry in genome-wide association studies (GWAS) is a critical issue that has significant implications, including hindering the progress of precision medicine initiatives. This issue is particularly significant in the context of neurodegenerative diseases (NDDs), where current therapeutic approaches have shown limited success. Addressing this under-representation is crucial to harnessing the full potential of genomic medicine in underserved communities and improving outcomes for NDD patients.

**Objective:** Our primary objective was to assess the representation of non-European ancestry participants in genetic discovery efforts related to NDDs. We aimed to quantify the extent of inclusion of diverse ancestry groups in NDD studies and determine the number of associated loci identified in more inclusive studies. Specifically, we sought to highlight the disparities in research efforts and outcomes between studies predominantly involving European ancestry participants and those deliberately targeting non-European or multi-ancestry populations across NDDs.

**Evidence Review:** We conducted a systematic review utilizing existing GWAS results and publications to assess the inclusion of diverse ancestry groups in neurodegeneration and neurogenetics studies. Our search encompassed studies published up to the end of 2022, with a focus on identifying research that deliberately included non-European or multi-ancestry cohorts. We employed rigorous methods for the inclusion of identified articles and quality assessment.

**Findings:** Our review identified a total of 123 NDD GWAS. Strikingly, 82% of these studies predominantly featured participants of European ancestry. Endeavors specifically targeting non-European or multi-ancestry populations across NDDs identified only 52 risk loci. This contrasts with predominantly European studies, which reported over 90 risk loci for a single disease.

Encouragingly, over 65% of these discoveries occurred in 2020 or later, indicating a recent increase in studies deliberately including non-European cohorts.

**Conclusions and relevance:** Our findings underscore the pressing need for increased diversity in neurodegenerative research. The significant under-representation of non-European ancestry participants in NDD GWAS limits our understanding of the genetic underpinnings of these diseases. To advance the field of neurodegenerative research and develop more effective therapies, it is imperative that future investigations prioritize and harness the genomic diversity present within and across global populations.

**Abstract and highlights:** *Key Points:* Question
What is the state of ancestral inclusivity in genetic studies of neurodegenerative diseases? Findings
A systematic review of 123 publications on neurodegenerative diseases shows a focus on European populations, with only 18% of studies including any non-European ancestry data. Among 52 novel loci identified in non-European studies, 28 were from multi-ancestry studies (which included Europeans), 21 from East Asian studies, and 3 from other populations. Meaning
This significant disparity underscores the need for more inclusive research approaches in neurodegenerative diseases, emphasizing multi-ancestry and non-European populations to advance precision medicine and develop treatments effective for diverse populations.

## Introduction

Genome-wide association studies (GWAS) have shown a significant bias towards individuals of European ancestry, despite comprising only 16% of the global population ^1^. This underrepresentation issue is particularly salient in the realm of neurodegenerative disease (NDD) studies. For instance, while a recent Alzheimer’s disease (AD) GWAS including ∼800,000 individuals of European descent identified 75 disease-associated loci ^2^, no GWAS studies on AD currently exist for Admixed American or Native American populations. Similarly, Parkinson’s disease research exhibits a glaring imbalance, with black individuals included in just ∼4% of published PD studies ^3^.

The prevalence of NDDs varies significantly among global populations and racial/ethnic groups. This warrants a critical examination of the disparity in genetic research efforts over time. In this manuscript, we present a systematic review spanning from 2012 through 2022, focusing on neurodegenerative disease GWAS research.

Our analysis encompasses common NDDs such as Alzheimer’s disease, Parkinson’s disease, and amyotrophic lateral sclerosis, as well as less common atypical dementias. Our objective is to quantify the disparity in participant recruitment for genetic studies, shed light on genetic findings in underrepresented populations, and discuss ongoing initiatives aimed at addressing this pervasive issue.

## Methods

### Search Strategy

The systematic review was conducted in two phases. First, we reviewed the <u>GWAS Catalog</u>; then, since the GWAS Catalog does not include all GWAS studies, we performed a formal literature review in collaboration with the National Library of Medicine (NLM). The keywords used in both searches are included in Supplementary Table 1.

Results from both the GWAS Catalog and the NLM search were uploaded to Covidence ^4^, a web-based software platform, for further review. We removed duplicate studies and any studies published before 2012 or after 2022.

All studies were filtered to only include genome-wide associations examining neurological disease risk factors, family history of disease, disease progression, age at onset, or survival genome-wide, excluding exome wide studies and those that focused on a targeted set of SNPs or genetic loci. Studies investigating disease subtypes, biomarkers, non-English language studies, and those investigating only rare or structural variation were also excluded.

Studies were assessed for eligibility by two independent reviewers and all conflicts were resolved by a third independent reviewer. Publication date, phenotype, and cohort information were extracted from each publication. If multiple phenotypes of interest were analyzed in the same study, information was included in both phenotype categories. The number of samples per ancestry was extracted manually from each study. We looked at seven ancestry groupings: European (EUR), East Asian (EAS), Middle-Eastern (MDE), African (AFR), African American and Caribbean (AAC), Latino and indigenous Americas populations (AMR), and South Asian (SAS). A PRISMA diagram of our filtering process can be found in Figure 1. All 123 studies passing our filters can be found in Supplementary Table 2.

**Figure 1:**
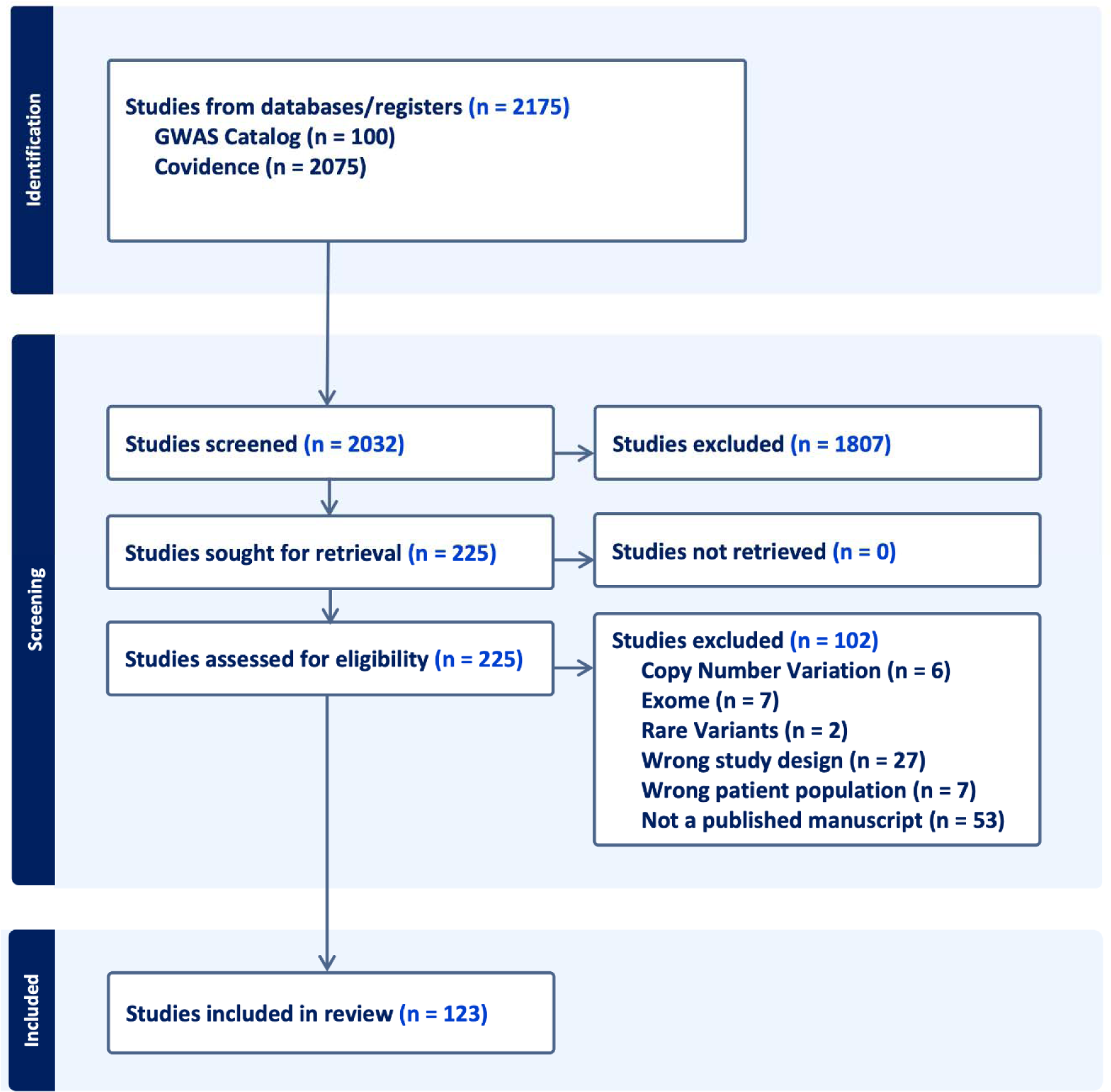
PRISMA flow diagram for systematic review of published GWAS studies and pubmed literature review from NLM. N in the figure relates to the number of published studies as of April 28th, 2022.

Finally, results were examined manually for all studies passing implemented filtering methods. Novel loci discovered in non-European or multi-ancestry populations, with a p-value below 5E-8, are listed in Table 2.

**Table 1:** Largest GWAS sample size by NDD and ancestry for single and multi ancestry studies.

| NDD | Ancestry | Author | Year | Total samples<br>(% non-European*) | Study |
| --- | --- | --- | --- | --- | --- |
| AD | AAC | Sherva | 2022 | 75,058 | African ancestry GWAS of dementia in a large military cohort identifies significant risk loci |
|  | EAS | Hirano | 2015 | 17,031 | A genome-wide association study of late-onset Alzheimer's disease in a Japanese population |
|  | EUR | Wightman | 2021 | 1,126,563 | A genome-wide association study with 1,126,563 individuals identifies new risk loci for Alzheimer's disease |
|  | MULTI | Lake | 2022 | 644,188<br>(2.83%) | Multi-ancestry meta-analysis and fine-mapping in Alzheimer's Disease |
|  | AMR,MDE |  |  | NO STUDIES |  |
| PD | AMR | Loesch | 2021 | 1,497 | Characterizing the Genetic Architecture of Parkinson's Disease in Latinos |
|  | EAS | Foo | 2017 | 14,006 | Genome-wide association study of Parkinson's disease in East Asians |
|  | EUR | Nalls | 2019 | 1,474,097 | Identification of novel risk loci, causal insights, and heritable risk for Parkinson's disease: a meta-analysis of genome-wide association studies |
|  | MULTI | Kim | 2022 | 2,525,730<br>(38.12%) | Multi-ancestry genome-wide meta-analysis in Parkinson's disease |
|  | AAC,MDE | NO STUDIES |  |  |  |
| ALS | EAS | Wei | 2019 | 4,727 | Identification of TYW3/CRYZ and FGD4 as susceptibility genes for amyotrophic lateral sclerosis |
|  | EUR | vanRheenen | 2016 | 41,398 | Genome-wide association analyses identify new risk variants and the genetic architecture of amyotrophic lateral sclerosis |
|  | MULTI | van Rheenen | 2021 | 152,268 (12.00%) | Common and rare variant association analyses in amyotrophic lateral sclerosis identify 15 risk loci with distinct genetic architectures and neuron-specific biology |
|  | AAC, AMR, MDE | NO STUDIES |  |  |  |
| MS | AAC | Isobe | 2015 | 2319 | An ImmunoChip study of multiple sclerosis risk in African Americans |
|  | AMR | Ordoñez | 2015 | 161 | Genomewide admixture study in Mexican Mestizos with multiple sclerosis |
|  | EUR | Patsopoulos | 2019 | 115,803 | Multiple sclerosis genomic map implicates peripheral immune cells and microglia in susceptibility |
|  | EAS,MDE, MULTI | NO STUDIES |  |  |  |
| FTD | EUR | Ferrari | 2014 | 12,928 | Frontotemporal dementia and its subtypes: a genome-wide association study |
|  | AAC, AMR, EAS, MDE, MULTI | NO STUDIES |  |  |  |
| MG | EAS | Na | 2014 | 259 | Whole-genome analysis in Korean patients with autoimmune myasthenia gravis |
|  | EUR | Chia | 2022 | 45,675 | Identification of genetic risk loci and prioritization of genes and pathways for myasthenia gravis: a genome-wide association study |
|  | MULTI | Sakaue | 2021 | 533,853 (33.48 %) | A cross-population atlas of genetic associations for 220 human phenotypes |
|  | AAC,AMR, MDE | NO STUDIES |  |  |  |
| LBD | EUR | Chia | 2021 | 7,372 | Genome sequencing analysis identifies new loci associated with Lewy body dementia and provides insights into its genetic architecture |
|  | AAC,AMR,EAS,MDE, MULTI | NO STUDIES |  |  |  |
| VaD | EUR | Moreno-Grau | 2019 | 4,830 | Genome-wide association analysis of dementia and its clinical endophenotypes reveal novel loci associated with Alzheimer's disease and three causality networks: The GR@ACE project |
|  | MULTI | Fongang | 2022 | 482,088 (2.40%) | A meta-analysis of genome-wide association studies identifies new genetic loci associated with all-cause and vascular dementia |
|  | AAC, AMR,EAS, MDE | NO STUDIES |  |  |  |

\*if applicable

**Table 2:** Genome-wide significant (P-value < 5E-8) novel loci nominated in non-European populations or multi-ancestry studies. We found no diverse or multi-ancestry loci for LBD, FTD, MG, or VaD. P-values are given in parentheses. Nominated loci were determined as the nearest gene or genomic context within 1MB of the significant SNP.

| Disease | Ancestry | Nominated loci |
| --- | --- | --- |
| AD | AAC | *COBL* (3.8E-8), *SLC10A2* (4.6E-8)<sup>11</sup> |
|  | EAS | *RHOBTB3/GLRX* (3.07E-19), *CTC-278L1.1* (2.49E-23), *CTD-2506J14.1* (1.35E-67), *CHODL* (4.81E-9)<sup>15</sup>, *FAM47E* (5.34E-9)<sup>16</sup>, *CACNA1A* (2.49E-8), *LRIG1* (1.51E-8)<sup>17</sup> |
|  | AMR | *FBXL7* (6.19E-09)<sup>13</sup> |
|  | MULTI | *OR2B2* (2.14E-8)<sup>16</sup>, *SORL1* (1.04E-8)<sup>18</sup>, *TRANK1* (3.49E-8), *VWA5B2* (3.75E-8)<sup>19</sup> |
| PD | EAS | *NDN/PWRN4* (3.14E-9)<sup>23</sup>, *RPL3* (2.72E-8)<sup>24</sup> |
|  | MULTI | *ITGA8* (1.3E-8)<sup>26</sup>, *SV2C* (1.17E-10)<sup>27</sup>, *MTF2* (1.15E-10), *RP11-360P21.2* (1.65E-10), *ADD1* (4.11E-9), *SYBU* (3.62E-9), *IRS2* (2.30E-9), *USP8:RP11-562A8.5* (6.45E-10), *PIGL* (2.93E-9), *FASN* (2.61E-9), *MYLK2* (3.86E-9), *AJ006998.2* (1.12E-9), *Y_RNA* (3.81E-9), *PPP6R2* (4.09E-10)<sup>7</sup>, *BST1* (4.41E-8)<sup>28</sup> |
| ALS | EAS | *CAMKIG* (2.92E-8), *CABIN1/SUSD2* (2.35E-9)<sup>31</sup>, *INPP5B* (2.24E-8), *IQCF5/IQCF1* (2.06E-9), *ITGA9* (2.55E-8), *PFKP* (2.46E-9), *MYO18B* (2.28E-10), *ALCAM* (4.00E-8), *OPCML* (8.43E-9), *GPR133* (8.45E-10)<sup>32</sup>, *TYW/CRYZ* (2.10E-14), *FGD4* (5.19E-9)<sup>33</sup> |
|  | MULTI | *GPX3-TNIP1* (1.3E-8)<sup>30</sup>, *ACSL5-ZDHHC6* (8.3E-9)<sup>34</sup>, *SOD1* (3.5E-18), *HLA* (3.5E-12), *SLC9A8-SPATA2* (3.2E-10), *ERGIC1* (5.6E-9), *NEK1* (6.9E-9), *COG3* (1.2E-8), and *PTPRN2* (1.8E-8)<sup>36</sup> |

## Results

### Search results

We identified **123** eligible GWAS studies. Unsurprisingly, we found that European populations were overrepresented in GWAS pertaining to NDDs (**Figure 2**). When non-European populations were included, the sample sizes were on average 15X smaller than the European ancestry samples included in the same disease category. The underrepresentation of non-European populations was particularly evident among the less common NDDs, including Lewy body dementia and frontotemporal dementia, where we did not identify any non-European or multi-ancestry GWAS studies using the outlined search methods. We have summarized the lack of diversity in genetic studies of NDDs in **Table 1**, and **Figure 2**.

**Figure 2:**
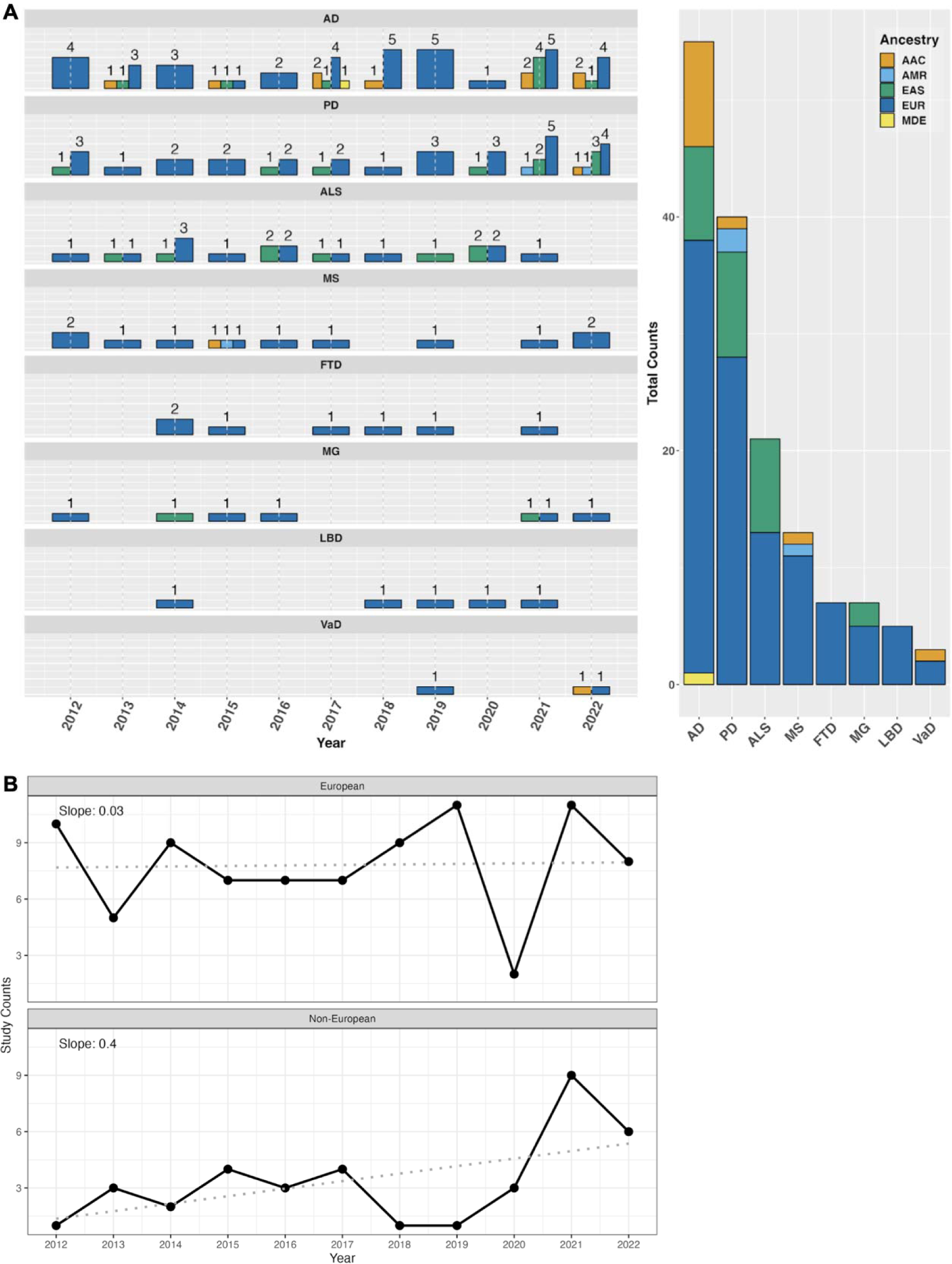
Number of studies over time from 2012 to 2022. A) Bar plot of study counts by NDD (left) with cumulative counts for each ancestry (right). Data in this figure includes both single and multi ancestry studies. B) Time series of the annual study counts in European and Non-European populations from 2012 to 2022. The slope from a linear regression is also displayed to highlight the rate of change in the number of study counts over time.

We found **52** novel NDD loci that were identified in non-European or multi-ancestry populations (**Table 2**). Of these **52** loci, **28** were found in multi-ancestry studies, **21** were found in East Asian studies, and only **3** were found in other populations (AAC and AMR). No loci were discovered in AFR, MDE, or SAS ancestries. Recent studies that combined individuals of multiple ancestries by using standard random-effects and some custom meta-analytic techniques ^5^ have succeeded in identifying novel disease loci that reach genome-wide significance, including two novel AD loci ^6^ and 12 novel PD loci ^7^. However, these studies leverage existing European sample sizes as a backbone for much of the statistical power needed for discovery.

In the following sections, we briefly summarize the results of our systematic review in a disease specific manner. Many of these findings have not been replicated. As datasets become larger and more inclusive, the genetic architecture of these diseases may grow and change.

### Alzheimer’s Disease

**Largest European GWAS**: Wightman 2021

**Total samples**: 1,126,563

**Largest multi-ancestry GWAS**: Lake 2022

**Total samples**: 644,188

**Total non-European samples:** 18,246

**% non-European:** 2.83%

**Largest non-European GWAS:** Sherva 2022

**Ancestry:** AAC

**Total samples**: 75,058

The largest Alzheimer’s disease GWAS of European populations included ∼1.1 million individuals and identified a total of 38 associated loci ^8^. Another recent GWAS included ∼800,000 individuals of European ancestry and identified a total of 75 loci ^2^. The discrepancy between identified loci in these studies could be due to many factors, including differences in neuropathological/diagnostic criteria ^9^.

A 2013 GWAS conducted in African Americans replicated an association at *ABCA7* previously identified in European populations. They found that rs115550680, rare in European populations, was associated with an increased risk for AD in African Americans comparable to the highly pathogenic APOE-D4 variant observed in Europeans ^10^. A 2017 GWAS in African Americans identified two novel loci at *COBL* and *SLC10A2* ^*11*^. The most extensive African American GWAS to date, drawing from a military cohort of around 22,000 individuals and a proxy GWAS involving approximately 50,000 individuals, identified significant associations with established AD risk genes such as *TREM2*, *CD2AP*, and *ABCA7*. Notably, distinct lead variants were observed in these loci compared to those found in European cohorts ^12^.

The only study conducted in Caribbean Hispanic individuals was a 2017 study with 2,451 cases and 2,063 controls. They found a novel and population specific locus near *FBXL7* ^*13*^. The lead SNP, rs75002042, is much more common in individuals with African ancestry compared to individuals of European ancestry, with minor allele frequencies around 20% and 0.009%, respectively. This study also replicated six loci previously reported in European populations, including *FRMD4A, CELF1, FERMT2, SLC24A4-RIN3, ABCA7,* and *CD33* ^13^.

The largest AD study in East Asian populations was conducted in Japanese participants with 1,827 cases and 15,204 controls (discovery + replication) ^14^, but they did not nominate any genome-wide significant loci. More recent but smaller studies have since been conducted, including a 2021 GWAS in a Chinese cohort that reported four novel loci near *RHOBTB3/GLRX, CTC-278L1.1, CTD-2506J14.1*, and *CHODL* ^*15*^, a study in Japanese participants that nominated a locus in *FAM47E* ^16^, and a study including both Korean and Japanese participants that nominated two novel loci at *CACNA1A* and *LRIG1* ^*17*^.

Multi-ancestry studies have nominated additional AD loci, however these studies still rely on Europeans as the majority population. *SORL1* was first identified as a risk locus for AD in a GWAS that included East Asian and European ancestry populations ^18^. Other multi-ancestry GWAS identified *OR2B2* ^*16*^, *TRANK1* and *VWA5B2* ^*19*^ as novel loci for AD.

While the inclusion of diverse populations in genetic research for AD is arguably better than what is seen for some of the atypical dementias, the largest study size for a non-European population ^12^ was still only 7% of the total sample size for the largest European AD GWAS.

### Parkinson’s disease

**Largest European GWAS**: Nalls 2019

**Total samples**: 1,456,306

**Largest multi-ancestry GWAS**: Kim 2022

**Total samples**: 2,525,730

**Total non-European samples:** 962,735

**% non-European:** 38.12%

**Largest non-European GWAS:** Foo 2017

**Ancestry:** EAS

**Total samples**: 14,006

The largest meta-GWAS of PD risk in individuals of European ancestry found 90 significant risk signals across 78 genomic regions. The 90 nominated risk variants collectively explain roughly 16-36% of the heritable risk of non-monogenic, or complex PD ^20^.

The largest study in East Asian populations (with exception to a study done in Japan before our review period ^21^) was conducted with Han Chinese participants, replicating loci previously identified in European populations including *SNCA*, *LRRK2* and *MCCC1* in their discovery GWAS of 14,006 participants ^22^. More recent studies in Chinese populations have nominated a locus on *NDN/PWRN4* associated with age at onset and a locus on *RPL3* associated with reduced survival ^23,24^.

The first and most recent PD GWAS of a South American population was conducted in 2021, replicating an association at *SNCA* with 1,497 participants ^25^.

Recently, more multi-ancestry studies have been conducted in PD, nominating novel loci for disease risk and age at onset including *ITGA8, SV2C, and BST1* ^*26*,27,28^. The largest meta-GWAS for PD, which included 4 ancestral populations, identified 12 novel loci: *MTF2, RP11-360P21.2, ADD1, SYBU, IRS2, USP8:RP11-562A8.5, PIGL, FASN, MYLK2, AJ006998.2, Y_RNA,* and *PPP6R2* ^*7*^.

The largest non-European PD GWAS was in East Asian populations, however, only a few novel loci have been nominated in that ancestry. Multi-ancestry studies have nominated more novel variants in recent studies, but much more work is needed to better understand risk for PD in non-European populations.

### Amyotrophic lateral sclerosis

**Largest European GWAS**: van Rheenen 2016

**Total samples**: 41,398

**Largest multi-ancestry GWAS**: van Rheenen 2021

**Total samples**: 152,268

**Total non-European samples:** 18,266

**% non-European:** 12.00%

**Largest non-European GWAS**: Wei 2019

**Ancestry:** EAS

**Total samples**: 4,727

ALS GWAS in European populations have nominated a number of risk loci including *C9ORF72*, *UNC13A*, *C21orf2*, *SARM1*, *MOBP*, *SCFD1, TBKK1,* and *KIF5A* ^29–30^.

ALS is the only disease in our review where more genome-wide significant novel loci have been identified in a non-European population than in the largest European-only study. The first GWAS of individuals with Chinese Han ancestry identified *CAMKIG* and *CABIN1/SUSD2* as susceptibility loci for ALS ^31^. Later studies in the Han Chinese population nominated additional novel loci including *INPP5B, IQCF5/IQCF1, ITGA9, PFKP, MYO18B, ALCAM, OPCML*, *GPR133, TYW/CRYZ* and *FGD4* ^32–33^. With a total of 12 genome-wide significant loci, East Asian ancestry GWAS for ALS have nominated the most of any single non-European population covered in our review.

Multi-ancestry GWAS for ALS, which typically consist of European and East Asian ancestry populations, have been successful at nominating additional risk loci including *GPX3-TNIP1* and *ACSL5* ^*30*,34,35^. The largest ALS GWAS to date was a multi-ancestry study including over 150,000 individuals of European and East Asian ancestry. This study identified a total of 15 risk loci for ALS, replicating 8 previously-identified and nominating 7 novel loci: *SOD1, HLA, SLC9A8-SPATA2, ERGIC1, NEK1, COG3,* and *PTPRN2* ^*36*^.

Similar to PD, ALS GWAS including or focused on East Asian populations have made more progress than other non-European populations for these diseases. However, much more work is still needed in all populations to progress potential precision medicine initiatives for ALS.

### Multiple Sclerosis

**Largest European GWAS**: Patsopoulos 2019

**Total samples**: 115,803

**There are no multi-ancestry studies in MS. Largest non-European GWAS:** Isobe 2015

**Ancestry:** AAC

**Total samples**: 2,319

The largest GWAS meta-analysis for MS included 115,803 individuals of European ancestry and found 82 significant genome-wide associations with MS. This study was also the first to identify a risk locus for MS on chromosome X and the identified genetic markers accounted for nearly 50% of the hereditary risk for MS ^37^.

Studies in non-European populations were more limited in MS than in the previous diseases discussed. The largest GWAS in African Americans was successful at replicating 21 of the loci previously identified in European populations, but did not nominate any new risk loci at a genome-wide significant level ^38^. The only other study nominated by our review process for MS in non-European populations was conducted in a Mexican population. The study found 4 significant variants, however, these variants had limited regional support and the study was severely underpowered with only 29 cases and 132 controls ^39^. Due to these limitations, we concluded that the variants identified in this study could not be classified as novel.

Sample sizes for MS GWAS are still relatively small, even for European populations. In addition, we did not find any multi-ancestry studies through our search methods, highlighting a potential opportunity for further discovery for this disease.

### Frontotemporal dementia

**Largest European GWAS**: Ferrari 2014

**Total samples**: 12,928

#### There are no multi-ancestry or non-European studies in FTD

Common risk loci nominated by previous European FTD studies include *C9ORF72, GRN,* and *MAPT* ^*40*^. The largest FTD GWAS in our review date range included ∼13,000 participants of European ancestry and nominated an additional locus in the *HLA-DRA/HLA-DRB5* region ^41^. This study was conducted in 2014, and while more recent GWAS of FTD have been performed, none have surpassed the sample size from the Ferrari study, and many have focused on smaller FTD subtypes ^42,43^.

No non-European or multi-ancestry GWAS were identified in our systematic review for FTD. Investigation of known genetic risk factors in non-Europeans suggest that *C9ORF72* expansions may be quite rare in Chinese populations,^44^ highlighting the need for further research in this area.

### Myasthenia Gravis

**Largest European GWAS**: Sakaue 2021

**Total samples**: 355,142

**Largest Multi-ancestry GWAS**: Sakaue 2021

**Total samples**: 533,853

**Total non-European samples:** 178,711

**% non-European:** 33.47%

**Largest non-European GWAS**: Na 2014

**Ancestry:** EAS

**Total samples**: 259

Known loci for MG include *PTPN22*, *CTLA4*, *HLA-DQA1, ZBTB10, and TNFRSF11A* ^*45*,46^, all nominated in European-based GWAS. The most recent GWAS for MG nominated an additional loci at *CHRNA1, SFMBT2,* and *FAM76B*, although the latter two did not replicate ^47^. The largest European and multi-ancestry GWAS for MG to date were both performed in the same study, leveraging 533,853 total samples from Japanese, UK, and Finnish-based biobanks. However, with only 278 cases, the effective sample size (4/(1/ncase+1/ncontrol)) for the meta analysis was insufficiently powered and they did not nominate any new loci for MG ^48^.

In non-European populations, the literature review identified one Korean GWAS for MG. However, this study was small and did not identify any loci meeting genome-wide significance ^49^. Other studies have found that there is earlier onset of MG in Asian populations, and higher prevalence of the ocular form in Asian children, highlighting the importance of continued discovery efforts for MG in non-European populations ^50^.

### Lewy body dementia

**Largest European GWAS**: Chia 2021

**Total samples**: 7,372

#### There are no multi-ancestry or non-European studies in LBD

Previously nominated risk loci for LBD include *GBA*, *APOE*, and *SNCA* ^51,52^. LBD can be hard to diagnose as there are a number of clinical and genetic overlaps with AD and PD, which may be one of the reasons why there is still limited genetic research for LBD in both European and non-European populations ^51,53^.

We found no LBD GWAS in any single non-European ancestry populations or any multi-ancestry studies through our search methods. Concrete data on the prevalence of LBD in diverse ancestries is difficult to acquire, showing a potential opportunity for valuable future research.

### Vascular dementia

**Largest European GWAS**: Moreno-Grau 2019

**Total samples**: 4,830

**Largest Multi-ancestry GWAS**: Fongang 2022

**Total samples**: 482,088

**Total non-European samples:** 11,590

**% non-European:** 2.40%

#### There are no non-European studies in VaD

Despite an approximated prevalence of about 15-20% in all dementia cases ^54^, vascular dementia remains difficult to study because of the uncertainty of diagnosis. In fact, only two studies on vascular dementia (VaD) passed our criteria and only one of these found genome-wide significant novel loci. The first study was a European GWAS that looked at vascular, mixed, and pure AD phenotypes and nominated loci at *ANKRD31* and *NDUFAF6* ^55^. The second study that passed our criteria was a multi-ancestry GWAS for all-cause and vascular dementia including participants from European, African, Asian, and Hispanic ancestries, but did not find any significant novel loci ^56^.

VaD prevalence and risk appears to be higher in South Asian ancestries compared to European or Chinese populations ^57,58^. Additional studies have suggested that African Americans are most likely to be admitted to inpatient care with a VaD primary diagnosis ^59^. Despite these findings, there are still limited genetic studies for VaD, and we found no single non-European GWAS, highlighting the need for future research.

## Discussion

This review highlights the lack of ancestral diversity in genetic research across neurodegenerative disease GWAS over the past decade. Current research suggests that including non-European populations can improve our understanding of the genetic architecture of disease through novel ancestry-specific discoveries, increased statistical power awarded by studying diverse haplotype structures, and the identification of loci with heterogeneous effects across populations ^63^.

Additionally, while we looked at seven genetic ancestry groups, these “buckets” do not capture the true diversity of global populations. The African continent is known to have high genetic diversity, yet individuals of African ancestry are routinely grouped into a single category ^64^. In fact, we found no studies investigating South Asian (SAS) or continental African (AFR) populations. After investigating the cohorts in our review, we noted that though there were multiple “African” labeled studies, none of them directly investigated individuals in *continental* Africa, instead looking at African-American or other African-admixed populations. It is critical to mention the reference population used to define the specific population, to prevent the misattribution of genetic features across ancestries. Grouping all participants with any African ancestry into a generalized African category obscures the significant issue of inadequate representation of continental Africans.

In addition, there has been very little research done on admixed populations, and how the combinations of different ancestries affect SNP frequencies and/or gene expression. A GWAS in a Caribbean Hispanic admixed population found that the frequency of a novel locus spanning *FBXL7* varied greatly, from 1% in those with European ancestry to 20% in African Americans.^13^

Furthermore, previous research has shown that the transferability of polygenic risk scores from African Americans to various African populations is highly unreliable ^65^. The substantial genetic and environmental disparities among individuals of African descent underscore the urgent need to improve diversity in genetic studies.

### Diversity in SNP discovery

While 6 of the 8 NDDs we investigated had non-European representation, only PD had >1,000 cases and >30% non-European samples (Table 1). Additionally, no new significant loci have been identified in diverse population studies for MS, LBD, FTD, VaD, or MG. While the largest non-European cohort in MG included almost 180,000 samples, only 81 MG cases were included. A GWAS with less than 1,000 cases is unlikely to achieve sufficient statistical power for SNP discovery in polygenic diseases where multiple loci with small effect sizes are generally expected ^60–62^. Alzheimer’s, the most well funded of all the NDDs, has less than 3% diversity among cases in genetic studies. The incorporation of studies that lack statistical power and replicability minimizes the true imbalance between European and non-European studies, maintaining a Eurocentric bias.

In addition, many genetic association studies in East Asian populations did not meet our review criteria because they were not conducted on a genome-wide scale. Instead, these studies often investigated only one or a small group of SNPs that had been previously associated with disease in European populations, potentially missing associations that are specific to non-European populations.

In fact, many loci identified in European populations have heterogeneous effects or ancestry-specific SNP associations. For example, while the *APOE* alleles account for around a quarter of overall heritability for AD in Europeans ^8,16^, several studies suggest that the *APOE4* allele has a weaker effect in African ancestry ^66,67^ and Caribbean Hispanic ^68^ populations. The effect has been found to be greater in Japanese populations ^66,67^. Heterogeneity of risk at *APOE4* has been quantified in a recent multi-ancestry meta-analysis, with an I2 up to 85%, with 50% or more of that risk heterogeneity attributable to genetic ancestry differences ^6^. We believe that examination of local ancestry at loci with such global differences may help discern whether locus-specific inheritance patterns modulate disease risk.

Similarly, *C9ORF72* is one of the most common risk factors for ALS. However, the frequency of the *C9ORF72* expansion is lower in Chinese populations (0.3%) as compared to European populations (7%) ^30^. Recent research suggests that commonly used genetic tests to diagnose ALS may be less accurate in non-European ancestry patients because they are less likely to carry the *C9ORF72* structural variant ^69^.

Some SNPs with large effect sizes don’t exist or are extremely rare in certain ancestry groups. Variants in *ABCA7*, for example, increase AD risk more in individuals of African ancestry than in those of European ancestry ^70^. In fact, *ABCA7* has a comparable effect size to *APOE* in individuals of African ancestry ^10^. Genetic variants in *LRRK2, GBA*, and *SNCA*, which have been associated with increased risk of PD in European ancestry populations, appear to have a negligible effect in individuals from India ^71,72,73,74^. Without studying diverse populations, researchers would miss the population-specific effects of these loci and potential therapeutic targets which modify their effects.

### Looking forward

Despite the inequalities highlighted above, progress is being made. Researchers in AD are taking a strong multi-modal approach to increasing diversity. The Multi-Partner Consortium to Expand Dementia Research in Latin America (<u>ReDLat</u>) is leveraging “on the ground” connections with research communities in Latin America and the Caribbean to grow a diverse database of dementia resources ^75,76^. The Alzheimer’s Disease NeuroImaging Study (<u>ADNI</u>) is growing more inclusive cell lines and generating partner data for multiple ancestrally diverse samples ^77^. The NIH’s Center for Alzheimer’s and Related Dementias (<u>CARD</u>) is filling diversity gaps by creating training materials, generating data to complement existing efforts, and providing open science support for researchers in diverse communities.

Multiple efforts are also underway in the PD space. The Genetic Architecture of Parkinson disease in India (GAP-India) plans to develop a large clinical/genomic biobank in India ^71^. The Latin American Research Consortium on the Genetics of PD (<u>LARGE-PD</u>) aims to address inclusivity and genomic differences within and across Latino populations. Finally, the Global Parkinson’s Genetics Program (<u>GP2</u>) aims to genotype >150,000 individuals from around the world. GP2-funded projects include the Black and African Americans Connections to Parkinson’s Disease Study (<u>BLAACPD</u>), which seeks to assess the genetic architecture of Black and African American individuals with Parkinson’s disease, as well as healthy subjects, from across the United States ^78^. GP2 is motivated to increase diversity not only just among samples recruited into studies, but also in the investigators making use of the data, providing training and resources to ensure that all researchers are on an open and equal field of play ^79^.

A list of ongoing efforts for increasing diversity in NDD genetic research, including atypical dementias, can be found in Supplemental Table 3.

These efforts are paying off. More than 65% of the neurodegenerative disease-associated loci discovered in non-European or multi-ancestry populations were identified in the period between 2020 to 2022 (Figure 3). Over the past 10 years, there has been a steady increase in the proportion of non-European samples included in genetic studies (Figure 2B). With the increase in diverse samples in recent years, there has also been a growing interest in the use of multi-ancestry analyses to discover, fine-map and assess heterogeneity at disease risk loci, particularly in AD, PD, and ALS (Figure 2A).

**Figure 3:**
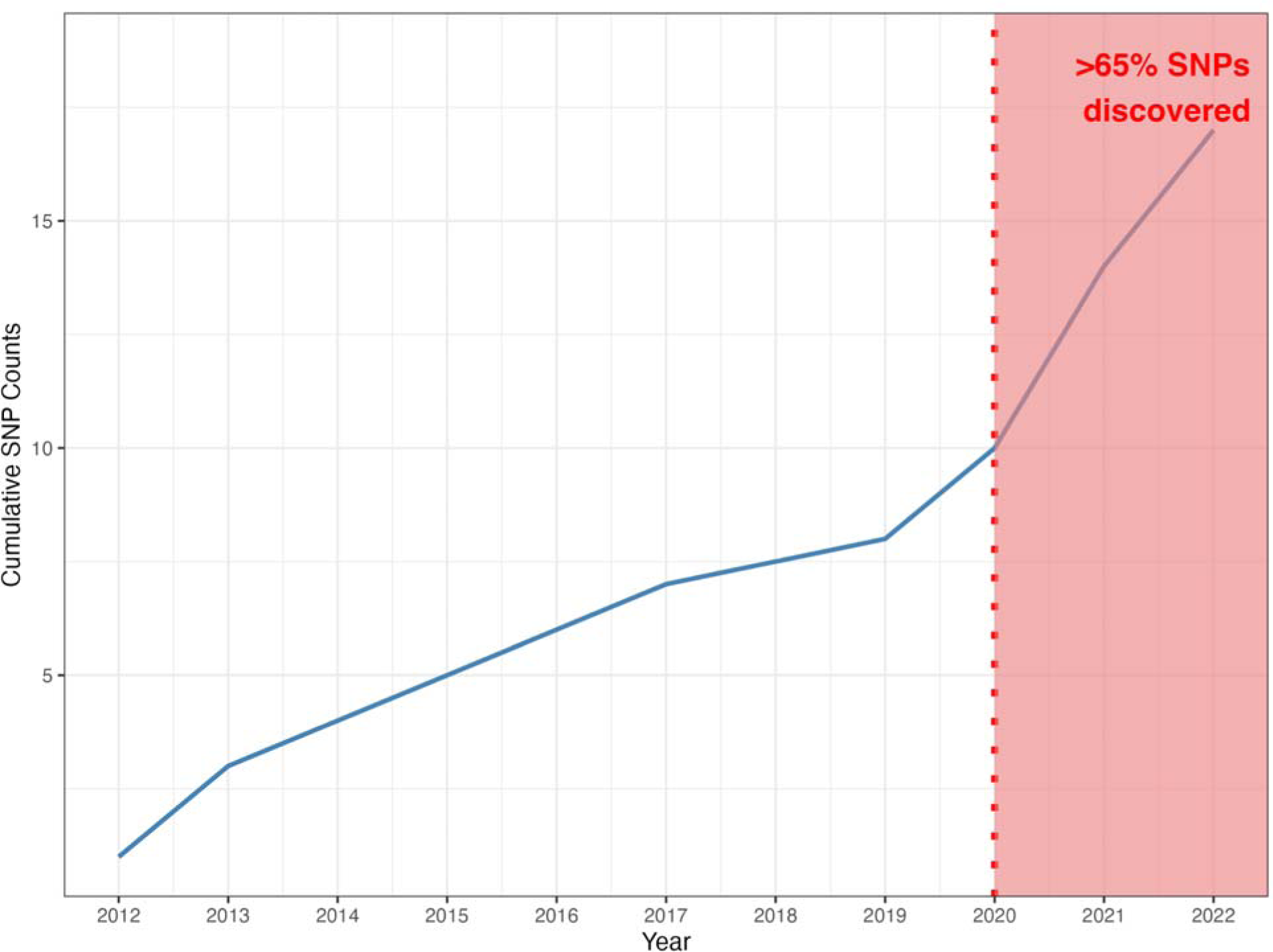
Cumulative count of discovered SNPs from 2012 through 2022. Notably, more than 65% of the SNPs were identified in the period between 2020 through 2022.

In fact, we are already seeing the benefits of increased diversity on genetic discovery in NDD research. A 2023 GWAS using African and African American samples collected by GP2 and 23andMe, and co-lead by researchers in Nigeria and NIH, found a novel *GBA1* locus that is rare in other populations^82^. We anticipate that in the future, leveraging multiple ancestries will continue to improve fine-mapping resolution to prioritize causal variants^5^, increase access to and reduce bias in precision medicine practices such as polygenic risk prediction^1^, and drive many new discoveries in the genetics of NDDs.

## Conclusion

Our systematic review highlights a striking disparity in the representation of diverse genetic ancestry populations in NDD research, emphasizing the urgent need for greater inclusivity to advance our understanding of these complex conditions and develop more equitable precision medicine approaches. Efforts to bridge this gap and promote diversity in genetic studies are vital for achieving meaningful progress in the diagnosis, treatment, and prevention of NDDs across global populations.

## Data Availability

All data produced in the present work are contained in the manuscript

## Acknowledgements

This research was supported in part by the Intramural Research Program of the NIH, National Institute on Aging (NIA), National Institutes of Health, Department of Health and Human Services; project number ZO1 AG000535, as well as the National Institute of Neurological Disorders and Stroke. We would also like to thank the National Library of Medicine, including Nancy Terry and Alicia Livinski, for their assistance with the review and Covidence. J.S.Y. is supported by NIH-NIA R01AG062588, R01AG057234, P30AG062422, P01AG019724, U19AG079774; NIH-NINDS U54NS123985; NIH-NIDA 75N95022C00031; the Rainwater Charitable Foundation; the Alzheimer’s Association; the Global Brain Health Institute; and the Mary Oakley Foundation.

